# The Risks and Benefits of Providing HIV Services during the COVID-19 Pandemic

**DOI:** 10.1101/2021.03.01.21252663

**Authors:** John Stover, Sherrie L. Kelly, Edinah Mudimu, Dylan Green, Tyler Smith, Isaac Taramusi, Loveleen Bansi-Matharu, Rowan Martin-Hughes, Andrew N. Phillips, Anna Bershteyn, for the HIV Modeling Consortium

## Abstract

**Introduction:** The COVID-19 pandemic has caused widespread disruptions including to health services. In the early response to the pandemic many countries restricted population movements and some health services were suspended or limited. In late 2020 and early 2021 some countries re-imposed restrictions. Health authorities need to balance the potential harms of additional SARS-CoV-2 transmission due to contacts associated with health services against the benefits of those services, including fewer new HIV infections and deaths. This paper examines these trade-offs for select HIV services.

**Methods:** We used four HIV simulation models (Goals, HIV Synthesis, Optima HIV and EMOD) to estimate the benefits of continuing HIV services in terms of fewer new HIV infections and deaths. We used three COVID-19 transmission models (Covasim, Cooper/Smith and a simple contact model) to estimate the additional deaths due to SARS-CoV-2 transmission among health workers and clients. We examined four HIV services: voluntary medical male circumcision, HIV diagnostic testing, viral load testing and programs to prevent mother-to-child transmission. We compared COVID-19 deaths in 2020 and 2021 with HIV deaths occurring now and over the next 50 years discounted to present value. The models were applied to countries with a range of HIV and COVID-19 epidemics.

**Results:** Maintaining these HIV services could lead to additional COVID-19 deaths of 0.002 to 0.15 per 10,000 clients. HIV-related deaths averted are estimated to be much larger, 19 - 146 discounted deaths per 10,000 clients.

**Discussion:** While there is some additional short-term risk of SARS-CoV-2 transmission associated with providing HIV services, the risk of additional COVID-19 deaths is at least 100 times less than the HIV deaths averted by those services. Ministries of Health need to take into account many factors in deciding when and how to offer essential health services during the COVID-19 pandemic. This work shows that the benefits of continuing key HIV services are far larger than the risks of additional SARS-CoV-2 transmission.

## Introduction

Many health services, including those providing HIV prevention, testing and care, have been restricted during the COVID-19 pandemic because services have been closed for some periods of time, capacity has been limited, or people were reluctant to access health services because of the risk of becoming infected. WHO released guidelines for essential health services that prioritized maintaining services for antiretroviral therapy (ART) [1]. Data reported to UNAIDS on monthly HIV service provision illustrates the extent of the problem [2]. These data show severe reductions in diagnostic testing, numbers of people newly initiating ART, viral load testing, programs to prevent mother-to-child transmission of HIV (PMTCT), services for key populations, voluntary medical male circumcision (VMMC), public sector condom distribution, and pre-exposure prophylaxis (PrEP). The severity of disruption varies by country and time but was as high as 75% in some cases. Service levels rebounded quickly to pre-COVID-19 levels for some services (PMTCT, key population services, VMMC, PrEP) in some countries but remained low through September 2020 for others. As of early 2021 increases in COVID-19 cases have led some countries to re-impose lock down measures.

For example, consider the situation in Zimbabwe. At the end of March 2020, Zimbabwe released guidance for several HIV services [3]. For VMMC, all road shows, sports galas and other events were suspended, traditional/religious circumcision camps were shuttered and VMMC services limited to walkins only. Guidance for HIV testing prioritized self-testing and limited facility-based testing to priority populations. For PrEP and ART multi-month dispensing was encouraged as well as efforts to limit crowding by revising visit schedules. Level 5 lockdown measures were re-imposed for 30 days in January 2021. These restrictions include restricted opening hours for essential services, closure for non-essential services, restrictions on attendance at funerals and weddings, limited public transportation services and staying at home except for buying food and medicines or transporting sick relatives. Similar restrictions were imposed in Malawi leading to only 33% of the expected VMMCs and large reductions in PrEP initiation and use.

Restricting some services makes sense if the benefits of those services are small compared to the potential harm of additional COVID-19 transmission. If the benefits are large, then there will be advantages to continuing those services even if it means some additional COVID-19 transmission and deaths. The purpose of this paper is to examine the benefits and disadvantages of providing HIV services during the COVID-19 pandemic.

## Methods

We used four HIV simulation models to estimate the HIV-related deaths that would be averted through delivery of HIV services had they been available and three COVID-19 simulation models to estimate the additional COVID-related deaths that could happen as a result of keeping the HIV services open. The COVID models looked at the possibilities of additional transmission from infected patients to health care workers and vice versa as well as potential transmission during patient travel to and from health clinics and thereafter within the household and beyond to the general population. COVID-19 and HIV were compared on the basis of additional COVID-19 deaths caused by continuing HIV services and HIV-related deaths averted. Since COVID-19 deaths would occur immediately and HIV-related deaths would occur later, possibly 30 years or more later if patients with new infections receive antiretroviral therapy (ART), we discounted all HIV-related deaths to the present year using a 3% annual discount rate as recommended for comparisons of this type [4]. Different models were applied to different countries with different levels of HIV and COVID-19 epidemics as described below. To ease comparisons across countries, all results are reported per 10,000 people receiving each HIV service.

The HIV services we examined included: the provision of voluntary medical male circumcision (VMMC) which reduces susceptibility to HIV transmission, HIV diagnostic testing which provides an entryway into treatment, viral load testing which determines whether ART is successfully controlling HIV, and programs to prevent mother-to-child transmission (PMTCT) which provides HIV testing and antiretroviral therapy to prevent transmission of HIV from mother to newborn. We did not include antiretroviral treatment in this analysis since previous work [5] has already shown the enormous benefits to continuing treatment services and most programs have followed guidelines and prioritized continuing these services. PrEP was not included because of the slow rollout of PrEP programs to date.

## HIV Models

The four HIV simulation models are described below. In each case, the team responsible for developing the model applied it to one or more countries using the best available data on HIV prevalence, behaviors and treatment over time.

### Goals Model

Goals is a compartmental HIV simulation model developed by Avenir Health [6]. It divides the adult population by sex and risk behaviors (not sexually active, one sexual partner in the last year, more than one sexual partner in the last year, female sex workers, male clients of sex workers, men who have sex with men and injecting drug users). HIV transmission occurs due to contact between an HIV-infected and HIV-uninfected individual. The probability of transmission is determined by the basic transmissibility of the virus and the characteristics of the contact, including the direction of transmission (male-to-female or female-to-male), the type of contact (heterosexual sex, male-male sex, needle sharing), stage of infection, presence of another sexually transmitted infection in either partner, condom use, male circumcision, viral suppression through ART, and use of pre-exposure prophylaxis (PrEP), the number of contacts per partner and the number of partners per year. Behavior change interventions affect transmission by changing risk behaviors, such as condom use, number of partners or age at first sex. People living with HIV are tracked over time by age, CD4 category and ART status. Mortality depends on age, CD4 count, and ART status. The model was applied to countries with different levels of HIV prevalence (India 0.22%, Kenya 4.5%, Malawi 8.9%, Nigeria 1.3%, South Africa 19% and Zimbabwe 12.8% in 2019) and COVID-19 cumulative cases reported per 10,000 population (India 75, Kenya 20, Malawi 4, Nigeria 4, South Africa 183 and Zimbabwe 10, as of December 2020). The model was used to estimate the impact of services for VMMC, condom distribution, HIV testing and PMTCT services.

### HIV Synthesis

The HIV Synthesis model is an individual-based model of sexual behaviour, transmission and progression of HIV and the effect of ART in an adult population updating in 3 month time steps. For each model run we sample from distributions of parameters and generate a range of “setting scenarios” which we consider reflect the diversity of epidemic and programmatic situations in sub-settings within countries sub-Saharan Africa, as well as reflecting uncertainty over some parameter values. The effect of opening three services is modelled: HIV testing, viral load monitoring and ART switching, and VMMC. We assume that each of these stopped on 1^st^ April 2020. We then consider 5 service resumption policies: (i) all three services remain unavailable until 1 July 2021; (ii) viral load monitoring and ART switching are resumed on 1 July 2020, HIV testing and VMMC remain unavailable until 1 July 2021; (iii) HIV testing is resumed on 1 July 2020; Viral load monitoring and ART switching and VMMC remain unavailable until 1 July 2021; (iv) HIV testing and Viral load monitoring and ART switching are resumed on 1 July 2020, VMMC remain unavailable until 1 July 2021; (v) all three services are resumed on 1 July 2020. We assume that people on ART are able to continue taking ART throughout this period (i.e., that this service is not disrupted), but the suspension of HIV testing impacts on ART initiations. The effect of resuming HIV testing on numbers of deaths per 10,000 people receiving the service is derived from the difference between policies (i) and (iii), the effect of resuming viral load monitoring from the difference between policies (i) and (ii), and the effect of resuming VMMC services from the difference between policies (iv) and (v).

### Optima HIV

The Optima HIV model is a compartmental model with populations disaggregated by sex, age, and risk. Models for 38 countries (Argentina, Armenia, Belarus, Brazil, Bulgaria, Cambodia, Colombia, Democratic Republic of the Congo, Eswatini, Ethiopia, Haiti, India, Iran, Kazakhstan, Kenya, Kyrgyzstan, Macedonia (Former Yugoslav Republic), Malawi, Mexico, Moldova, Mozambique, Nepal, Nigeria, Papua New Guinea, Peru, Russian Federation, Senegal, South Africa, Sudan, Tajikistan, Tanzania, Thailand, Togo, Uganda, Ukraine, Uzbekistan, Vietnam, and Zimbabwe) were used in this analysis. The effect of opening each HIV service was modelled in countries based on where HIV services were implemented in a comparable format. This included nine countries in sub-Saharan Africa with coverage disaggregated by VMMC and traditional circumcision, 19 countries with viral load monitoring coverage disaggregated from ART, and all 38 countries for HIV testing. ART and PMTCT coverage were held constant over the study period to reflect 2020 values for the proportion of people diagnosed with HIV, while coverage of other programs was held constant based on the latest reported spending.

The same analysis approach described above for the HIV Synthesis model was used. The number of client visits averted by not re-opening services was based on the assumption that each HIV test, each viral load test, and each voluntary medical male circumcision consisted of one client visit, giving the deaths averted per 10,000 clients as 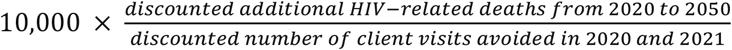. Models were run independently over 50 years with reopening of each service on 1 July 2020 compared with 1 July 2021. A 2.4-month time step was applied, with a resulting 5-timesteps per year. Mean estimates are reported (weighted by the number covered by each service for each country) with uncertainty given as the 25^th^ and 75^th^ percentile for individual countries to capture the variation across countries representing local conditions.

### EMOD

EMOD-HIV is a network transmission model of HIV [7] built into the open-source individual-based modeling framework EMOD [8]. HIV transmission is simulated at the level of each coital act between sexual partners, who form an age/sex-structured network of relationships of variable duration [9]. The model was fit to epidemic patterns in South Africa including programmatic data on HIV treatment and survey estimates of age/sex-specific HIV incidence and prevalence by adjusting parameters related to sexual risk behavior and engagement in care, as described elsewhere [10-12]. Age/sex-specific risk of non-HIV deaths were applied to the population based on demographic projections from the United Nations Population Division, and a competing risk of HIV-associated death was applied to PLHIV on ART and not on ART. For PLHIV not on ART, risk of HIV-associated death depends on age at time of infection, with older adults progressing more rapidly toward AIDS and death. For PLHIV on ART, risk of HIV-associated death depends on age, sex, and CD4 count at time of ART initiation and level of ART adherence, with declining risk of death on ART for adherent patients who remain stable on ART [13]. For individuals who interrupt and resume ART, risk of death calculated based on CD4 count at time of interruption and re-engagement in ART [14]. We compared the number of HIV-associated deaths in model projections with different durations of service disruption, as described above for the HIV Synthesis model, applied to the services of HIV testing, VMMC, and PMTCT. The number of interventions delivered was calculated in order to obtain the ratio of the difference in the number of deaths to the difference in the number of clients served, and the ratio was multiplied by 10,000 in order to obtain the reported results. Because EMOD-HIV is a stochastic model, simulations were run 250 times and 95% uncertainty intervals were reported, with the constraint that delivery of the services could not increase the number of HIV-associated deaths due to stochastic effects.

## COVID-19 Models

Three COVID-19 simulation models were applied to estimate the potential COVID-related deaths that could occur due to transmission related to HIV services.

### Contact Model

We developed a simple contact model of COVID-19 transmission based on an approach developed by Abbas et al. to examine the risks related to childhood immunization. [15] We estimate potential new infections among health care workers, clients and the general public. New infections among health care workers *I*_*HW*_ are estimated as:

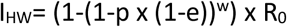

Where *p* is the prevalence of infectious SARS-CoV-2 in the population, *e* is the effectiveness of personal protective equipment used by health care workers (set at 90%), *w* is the number of health care workers that have contact with each client and R_0_ is the average number of people who will contract an infection from one infected person (which accounts for onwards transmission from the health care worker).

Client infections at health care settings are estimated as:

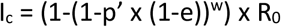

where *p’* is the prevalence of infections SARS-CoV-2 in health care workers. This is assumed to be lower than the general population due to the use of personal protective equipment.

For client contact with the general public during travel to and from, and while awaiting, services the probability of infections is estimated as:

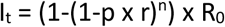

where *r* is the probability of infection given contact with an infectious person during travel or in the waiting area and *n* is the number of contacts during travel to and from the clinic or in the clinic waiting area.

The population prevalence of infectious SARS-CoV-2 is estimated as the average reported number of daily new cases of COVID-19 multiplied by the average duration of the infectious period (in days) divided by the adult population size. While new daily cases are likely to be under-reported [16] leading to an under-estimate of new COVID-19 cases, we also calculate the COVID-19 mortality rate from the reported COVID-19 cases and deaths so that the two measures will be consistent.

The number of deaths due to COVID-19 is estimated as the number of new infections multiplied by the mortality rate and adjusted for the age. The majority of COVID-19 deaths occur to people over the age of 65. Health care workers and clients are likely to be much younger. We adjust the population mortality rate downward to account for these age effects. In the United States the age group 35-44 accounts for 1.8% of all COVID-19 [17] deaths and 16.4% of COVID-19 cases [18], so the population mortality is multiplied by 0.11 to account for the age effects.

### Cooper/Smith COVID Model

We developed a deterministic, compartmental, age-structured transmission model for COVID-19 down to the sub-district level in Malawi beginning on April 1^st^ for one year. The model has a SEIR structure and estimates infections, hospitalization need, intensive care need, and deaths and includes daily estimates of cloth mask utilization and physical distancing. We adjust the risk of severe disease resulting in hospitalization, intensive care, and death according to age using age-specific rates from the literature. Finally, community spread of COVID-19 did not begin on the same date in each sub-district. In lieu of sufficiently granular testing data, we estimate each sub-district’s epidemic start date by leveraging estimates of connectedness between sub-districts from mobile phone data. These daily estimates of connectedness act as a likelihood of transmission based on the pool of susceptibles and infectious individuals in source and sink sub-districts, respectively.

Surveillance of infections, hospitalizations, and deaths are highly limited in Malawi, with one of the lowest testing rates in the world and very limited death registration. We triangulate limited seroprevalence estimates in Malawi, with seroprevalence estimates from other settings including South Africa in combination with their respective test rates. We estimate that actual infections of COVID-19 in Malawi may be undercounted by 500-times, while COVID deaths are undercounted by approximately times. We adjusted model parameters (within sensible range of literature review estimates) including R0, generation time, and hospitalization risk to align the projection with the calibrated trajectory of infections and deaths.

Over one year, we estimate approximately 3,500,000 COVID-19 infections and 6,600 deaths among adults. We then estimate what proportion of infections and deaths are attributable to HIV service delivery as those services are a proportion of total close contacts with similar transmission probability. We estimate under a range of HIV service delivery scenarios that those services contribute approximately 1.2%-2.5% of total contacts – and therefore that same proportion of infections and deaths. We express these COVID-19 infections and deaths as a rate per 10,000 HIV clients, and break results down by service type.

### Covasim

Covasim is an agent-based, age-and sex-structured SEIR model for COVID-19 transmission. A model for KwaZulu-Natal, South Africa was informed using estimates for household size distribution and contact network matrices (for home, school, workplace, and the community by 5-year age bands) and was calibrated to daily numbers of COVID-19 cases, deaths and tests using changes in viral transmission to reflect implementation of different COVID-19 policies over time. The model was also informed by global COVID-19 disease parameter estimates as described by Kerr et [19]. To simulate conditions used in the Contact model described above, contact with two (HIV, viral load testing) or three (VMMC, PMTCT) healthcare workers were specified and the probability of transmission per contact with an infectious member in the community was set to 6%. Transmission was simulated for 1-year from 1 March 2020. SARS-CoV-2 infections and COVID-19 deaths as a result of VMMC, HIV testing, viral load testing, or PMTCT service-related exposure were estimated and expressed per 10,000 HIV clients. A ratio of COVID-19 mortality for those aged 35-44 years to the total population of 0.11 was assumed following the Contact model approach.

## Results

Our simulations estimate that the additional COVID-19 deaths per 10,000 in-person HIV service clients ranges from 0.002 to 0.155 while the additional HIV-related deaths averted by keeping those services open range from 19 to 146. The average ratio of discounted HIV-related deaths to COVID-19 deaths ranges from 140 for HIV testing to 740 for VMMC services. Table 1 shows the detailed results by model and service.

**Table 1.** COVID-19 deaths among health care workers, clients and family members due to transmission during access to HIV services and HIV-related deaths that could be averted by these services per 10,000 clients

| HIV Service | Additional COVID-19 Deaths |  |  | HIV-Related Deaths Averted |  |  |  |
| --- | --- | --- | --- | --- | --- | --- | --- |
|  | Contact Model | Cooper/Smith | Covasim | Goals | HIV Synthesis | Optima HIV | EMOD |
| VMMC | 0.01-0.24 | 0.004 | 0.098 | 19<br>(10-30) | 146<br>(31-323) | 103<br>(4-223) | 92<br>(0-323) |
| HIV testing | 0.00-0.17 | 0.152 | 0.004 | 22<br>(20-35) | 20<br>(1-52) | 22<br>(12-42) | 14<br>(6-22) |
| Viral load testing | 0.00-0.17 | 0.152 | 0.004 |  | 44<br>(27-61) | 49<br>(39-275) |  |
| PMTCT | 0.01-0.24 | 0.002 | 0.098 | 33<br>(0-100) |  |  | 44<br>(0-358) |

Note: Figures in parentheses represent 95% confidence bounds. Ranges for Contact Model are across 6 countries. Different models were applied to different countries as specified in methods section. The rate of mortality is the unweighted averaged across the countries for which the analysis was conducted.

## Discussion

Our results clearly show that the potential increase in HIV transmission and subsequent death that would be associated with curtailing HIV services during the COVID-19 pandemic are many times larger than the expected COVID-19 deaths due to transmission during visits to access HIV services. COVID-19 deaths would have to be at least 1000 times higher than we estimated to result in an equivalent number of deaths.

As indicated by the ranges for the HIV outcomes in Table 1 these results are robust to uncertainties in model estimates. However, the results are sensitive to assumptions regarding the annual discount rate and the time from HIV-infection until HIV-related death. Without ART the median survival with HIV is about 11 years. With a 3% discounting rate, an additional death that occurs in 2031 (consistent with the 11-year median HIV survival time in the absence of treatment) would be discounted by a factor of 1.4 while an additional death that is postponed to 2050 due to treatment would be discounted by a factor of 2.5. A somewhat higher discount rate may be appropriate for many developing countries.[20] However, the discount rate would need to be as high as 18% to bring the discounted HIV down to the level of the COVID-19 deaths.

This result derives mostly from the differential mortality rates of the two diseases. HIV generally leads to death although a person with drug sensitive virus who takes their regimen every day is unlikely to die from HIV, even after 30 or 40 years [21]. If HIV is controlled long enough, a person living with HIV may die from some other cause before dying of HIV. In comparison, COVID-19 is rarely fatal. By the end of 2020 85 million cases and 1.8 million deaths had been reported globally, implying a mortality rate of about 2%. Although deaths are certainly under reported, the number of cases have an even larger under count. Most COVID-19 deaths are in older populations and in those with comorbidities. The mortality of people in their 30s and 40s may be about 0.2%. Of course, both COVID-19 and HIV also produce disabilities, besides. Only HIV-associated disabilities were considered in this study. Long-term disabilities among survivors of COVID-19 are not yet well-characterized due to the novelty of the virus [22]

The results presented here come from seven different models applied to different sets of countries and settings. The variation in the results largely reflect those differences. There are also differences in model structure relating to how age and risk behaviors are handled that affect the results. The modeling teams often made different assumptions about how to interpret available data and which dynamics were important to model explicitly. The models agree surprisingly well on some results and vary by an order of magnitude on others, which we hope to explore further in future collaborations. But the variation in estimates across models is quite small compared to the differences in estimated COVID-19-related and HIV-related mortality.

The contact model does not account for potential immunity to SARS-CoV-2 that may have accrued among health care workers and patients prior to potential exposures related to HIV service delivery. Thus, this model is expected to produce much higher COVID-19 risk than one that accounts for accrual of immunity, and therefore should be interpreted as an upper bound to COVID-19 risk. However, this upper bound is worth considering given uncertainties about the cumulative number of SARS-CoV-2 infections in these settings, the duration and effectiveness of SARS-CoV-2 immunity in these settings, and the potential for new variants of SARS-CoV-2 to evade immune responses.

There are limitations to this analysis. COVID-19 is a new disease, so models are also new and relatively untested in projecting into the future. We have modelled HIV services in isolation but they are imbedded within a larger health system and effects of shortages of health personnel to deal with COVID-19 hospitalizations, testing and vaccination campaigns may also affect HIV services. Decision to continue a specific HIV service, such as testing, may be determined more by decisions about how to provide all types of health services rather than just those related to HIV. Disruptions to supply chains may also affect the delivery of health services.

Most programs have taken steps to ensure that essential HIV services such as ART re-supply can continue. Often this involves multi-month scripting to reduce the number of visits required for patients doing well on ART. Countries have learned from past epidemics, such as Ebola, how to maintain essential services under extraordinary circumstances. [23]

Many health systems are under great strain trying to cope with the surge of hospitalizations due to COVID-19 and meanwhile maintain other essential health services, but many are finding ways to continue these important services in spite of the obstacles. These efforts are important not only for averting new HIV infections but also for averting other infectious diseases through continued childhood immunization, avoiding unintended pregnancies by continuing family planning services, avoiding increased TB transmission by continuing diagnoses and treatment programs, and for many other health challenges as well. Our efforts should focus on how to maintain these services so that we do not lose the benefits of the progress in HIV that has been made in recent years.

## Data Availability

All data are publicly available as indicated or provided in the article itself.

## Funding

JS received funding from the Bill & Melinda Gates Foundation (grant OPP1118702). ANP, LBM and AB received support from the Bill & Melinda Gates Foundation under grant OPP1191655 (HIV Modelling Consortium). SLK and RMH received funding from NHMRC under grant GNT1184616. TS and DG received financial support from the Bill & Melinda Gates foundation under grant OPP1134992 and the United Nations Foundation Digital Impact Alliance under grant UNF-20-1113.

## Competing Interests

No competing interests were disclosed.

## Notes

### Competing Interest Statement

The authors have declared no competing interest.

### Author Declarations

Study used only published secondary data. No IRB or ethics approval was required.

### Summary of Updates

Corrected the spelling of the last name for Sherrie L. Kelly

